# Prior sleep-wake behavior predicts mental health resilience among adults in the United States during the COVID-19 pandemic

**DOI:** 10.1101/2021.06.15.21258983

**Authors:** Mark É. Czeisler, Emily R. Capodilupo, Matthew D. Weaver, Charles A. Czeisler, Mark E. Howard, Shantha M.W. Rajaratnam

## Abstract

Rigorous nonpharmaceutical interventions (e.g., stay-at-home orders, remote-work directives) were implemented in early 2020 for coronavirus disease 2019 (COVID-19) pandemic containment in the U.S. During this time, increased sleep duration and delayed sleep timing were reported through surveys (Leone et al., 2021) and wearable data (Rezaei and Grandner, 2021), as were elevated adverse mental health symptom (Czeisler et al., 2020). Inter-relationships between sleep and mental health have not been examined using longitudinal objective sleep-wake data, during these abruptly imposed lifestyle changes.

We examined objective sleep-wake data and surveyed mental health data collected among 4,912 U.S. adult users of a validated sleep wearable (WHOOP, Boston, Massachusetts) before and during the COVID-19 pandemic. Comparing the pre-pandemic (January 1 to March 12, 2020) and acute pandemic-onset intervals (March 13 to April 12, 2020), participants exhibited increased mean sleep duration (0.25h [95% CI = 0.237-0.270]), later sleep onset (18m [17.378-20.045]) and offset (36m [35.111-38.106]), and increased consistency of sleep timing (3.51 [3.295-3.728] out of 100); all P < 0.0001. Generally, participants with persistent sleep deficiency and low sleep consistency had higher odds of symptoms of anxiety or depression, burnout, and new or increased substance use during the pandemic. Decreases in sleep duration (adjusted odds ratio [aOR] = 1.30, 95% CI = 1.03-1.65, P = 0.025) and sleep consistency (2.05 [1.17-3.67], P = 0.009) were associated with increased anxiety and depression symptoms during the pandemic. We suggest that sleep duration and consistency may be important predictors of risk of adverse mental health outcomes during a pandemic.

M.J. Leone, M. Sigman, D.A. Golombek. Effects of lockdown on human sleep and chronotype during the COVID-19 pandemic. *Curr Biol* **30**(16), R930–R931 (2020).

N. Rezaei N, M.A. Grandner. Changes in sleep duration, timing, and variability during the COVID-19 pandemic: Large-scale Fitbit data from 6 major US cities. *Sleep Health* 10.1016/j.sleh.2021.02.008. (2021).

M.É. Czeisler, R.I. Lane, E. Petrosky, et al., Mental Health, Substance Use, and Suicidal Ideation During the COVID-19 Pandemic - United States, June 24-30, 2020. *MMWR Morb Mortal Wkly Rep* **69**(32), 1049–1057 (2020).

**Significance Statement:** The coronavirus disease 2019 (COVID-19) pandemic has had profound effects on health, including increased sleep duration and worsened mental health. We examined associations between (1) objective sleep-wake data before and during the COVID-19 pandemic and (2) adverse mental health symptoms and substance use among users of a validated sleep wearable. We found that, in general, participants with persistent sleep deficiency and low sleep consistency had higher odds of symptoms of anxiety or depression, new or increased substance use, and burnout. Our findings suggest that sleep of sufficient duration and consistent timing are associated with mental health resilience, exemplified in this case by the impact of the pandemic and related abrupt lifestyle changes on adverse mental health symptoms.

## Introduction

Absent widespread testing or safe and effective coronavirus disease 2019 (COVID-19) vaccines in early 2020, stringent mitigation policies were implemented in the United States and around the world to contain transmission of severe acute respiratory syndrome coronavirus 2 (SARS-CoV-2), the virus that causes COVID-19. Among the many consequences of these measures was an enhanced opportunity for self-selection of sleep-wake timing, resulting from work-at-home directives, reduced travel and commutes, school closures, and stay-at-home orders. Survey data (1-7) and longitudinal wearable or mobile application data (8-12) have been used to report increased sleep duration and delayed sleep timing during this interval, in the U.S. and other countries. Simultaneously, population-level surveillance studies revealed considerably elevated levels of adverse mental health symptoms and substance use among U.S. adults, including three-to four-fold times the prevalence of anxiety and depression symptoms and two times the prevalence of suicidal ideation in the second quarter of 2020 compared to that of 2019 (13-15).

Prior to the pandemic, associations between impaired sleep (e.g., sleep disturbance and insufficient sleep) and adverse mental and behavioral health symptoms (e.g., anxiety symptoms, depression symptoms, substance use, and suicidal ideation) were well established (16). In anticipation of mental health challenges in the wake of the onset of the COVID-19 pandemic and stringent mitigation measures, characterizing optimal sleep-wake structures for a mentally healthy life was among multidisciplinary research priorities posed to minimize adverse mental health effects of the pandemic (17). Indeed, during the initial phase of the COVID-19 pandemic, links between poor-quality and insufficient sleep and adverse mental and behavioral health symptoms were reported based on survey data (5, 18-20). However, self-reported sleep-wake data may be subject to various biases (21-24). Moreover, these studies did not include measures of variability in sleep timing, for which emerging evidence has identified associations with depressed mood (25-28) and other adverse health outcomes (29-31).

In the present study, we examined objective sleep and mental health among U.S. users of a validated sleep wearable (WHOOP, Boston, Massachusetts) (32, 33) before and during the COVID-19 pandemic using a comprehensive set of sleep variables (duration, sleep onset, sleep offset, consistency of sleep timing, and wake after sleep onset). Our findings align with data from other sources (1-12) demonstrating on average increased sleep duration and delayed sleep timing during the pandemic, and extend the existing literature by revealing increased consistency of sleep timing. Moreover, we found that participants with persistent sleep deficiency and low sleep consistency had higher odds of adverse mental health symptoms, and that anxiety or depression symptoms were associated with decreased sleep duration from moderate (6-7h) to short (<6h) and decreased consistency of sleep timing from moderate (70-80 out of 100) to low (<70). These findings support further investigations of sleep as a modifiable risk factor that could be targeted by behavioral interventions designed to enhance mental health resilience.

## Results

During June 24-30, 2020, 20,717 of 139,237 eligible invited active U.S. WHOOP users aged ≥18 years completed Internet-based surveys (response rate = 14.9%, Fig. S1). Overall, 4,912 (23.7%) participants had nocturnal sleep episodes recorded for ≥70% of nights throughout each of three intervals (pre-pandemic [January 1 to March 12, 2020]; acute pandemic onset [March 13 to April 12, 2020]; early chronic pandemic [April 13 to June 30, 2020]). Of these, 3,845 (78.3%) also completed the 4-item Patient Health Questionnaire to screen for symptoms of anxiety and depression (34). The sample comprised 3,471 (70.7%) male and 3,802 (77.2%) non-Hispanic White (White) adults. Most participants were highly educated (n = 4,105 [83.6%] college-educated), employed (n = 4,417 [89.9%]), and reported high household income (e.g., ≥USD$100,000, n = 3,126 [65.5%]). Mean age was 39.7 ± 11.24 years (Table S1). See Fig. S1 for the survey flow and Table S1 for detailed participant characteristics.

Overall, compared to the 6.95 ± 0.687 h mean sleep duration in the pre-pandemic interval, mean sleep duration was 0.25 h (95% CI = 0.237 to 0.270, p < 0.0001) longer in the acute pandemic interval, and 0.09 h (95% CI = 0.076 to 0.107, p < 0.0001) longer in the early chronic pandemic interval (Fig. 1A, Table S2). In the overall sample, mean sleep duration remained significantly longer on weekend nights compared with weeknights (except for holidays), though the magnitude of difference dampened with time (Fig. 1A). Sleep consistency (scored on a 0 to 100 scale), which was generally lower on weekend nights compared to weeknights, increased during both COVID intervals compared to the pre-pandemic interval, increased by 3.51 points (95% CI = 3.295 to 3.728 p < 0.0001) in the acute pandemic interval, and 4.06 points (95% CI = 3.856 to 4.267, p < 0.0001) in the early chronic pandemic interval (Fig. 1B, Table S2). Wake after sleep onset (WASO) decreased by 0.05 h (95% CI = 0.031 to 0.074, p < 0.0001) in the acute pandemic interval compared to the pre-pandemic interval but did not between the early chronic pandemic and pre-pandemic intervals (difference = 0.01 h, 95% CI = -0.022 to -0.045, p > 0.99) (Fig. 1C, Table S2). Finally, sleep timing abruptly shifted to a later time (i.e., delayed) immediately following the declaration of the pandemic by the World Health Organization (WHO) on March 12, 2020, and over the course of the month, mean sleep onset was 18 m later (95% CI = 17.384 to 20.045, p < 0.0001) and sleep offset was 36 m later (95% CI = 35.111 to 38.106, p < 0.0001) compared to the pre-pandemic interval (Fig. 1D, Table S2). The delay in sleep onset was sustained throughout the early chronic pandemic interval (17 m [95% CI = 16.470 to 19.289, p < 0.0001]), while the delay in sleep offset attenuated to 25 m during that time (95% CI = 23.629 to 26.714, p < 0.0001).

**Fig. 1.**
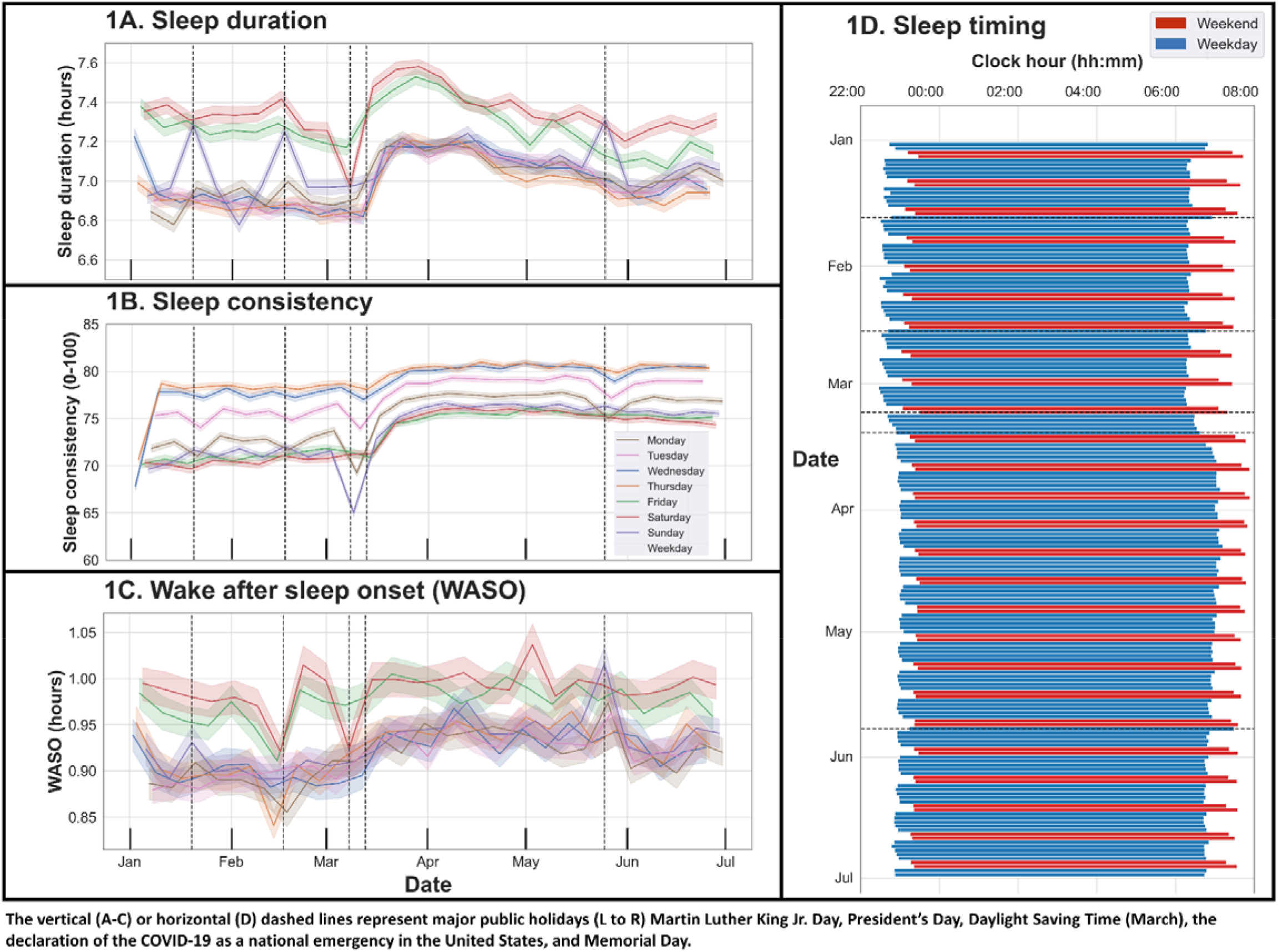
Sleep duration, consistency, wake after sleep onset, and timing, January 1, 2020—June 30, 2020.

While in the overall sample we observed longer sleep duration, increased consistency of sleep timing, relatively unchanged WASO, and delayed timing during the COVID-19 pandemic intervals, a subset of participants experienced marked changes in the opposite directions (Fig. 2). We therefore examined deciles of participants (n = 491) with the highest-magnitude changes in sleep duration, consistency, and WASO. The deciles with the highest-magnitude changes in sleep duration were lengthened or shortened by 0.77 h (95% CI = 0.742 to 0.794, p < 0.0001) and 0.50 h (95% CI = 0.522 to 0.470, p < 0.0001), respectively, while the deciles with the highest-magnitude changes in sleep consistency were increased and decreased by 12.85 points (95% CI = 12.480 to 13.214, p < 0.0001) and 4.41 points (95% CI = 4.720 to 4.099, p < 0.0001), respectively (Table S3). Regarding sleep timing, the deciles with the largest delays in sleep onset and offset shifted by 1 h 21 m (22:57 to 00:18, 95% CI = 77.284 to 84.861 m, p < 0.0001) and 1 h 39 m, (06:40 to 08:19, 95% CI = 95.456 to 102.811 m, p < 0.0001) respectively. The deciles with the largest advances in sleep onset and offset shifted by 33 m (23:29 to 22:55, 95% CI = 30.942 to 36.385 m, p < 0.0001) and 28 m (07:12 to 06:43, 95% CI = 26.036 to 31.358 m, p < 0.0001), respectively.

**Fig. 2.**
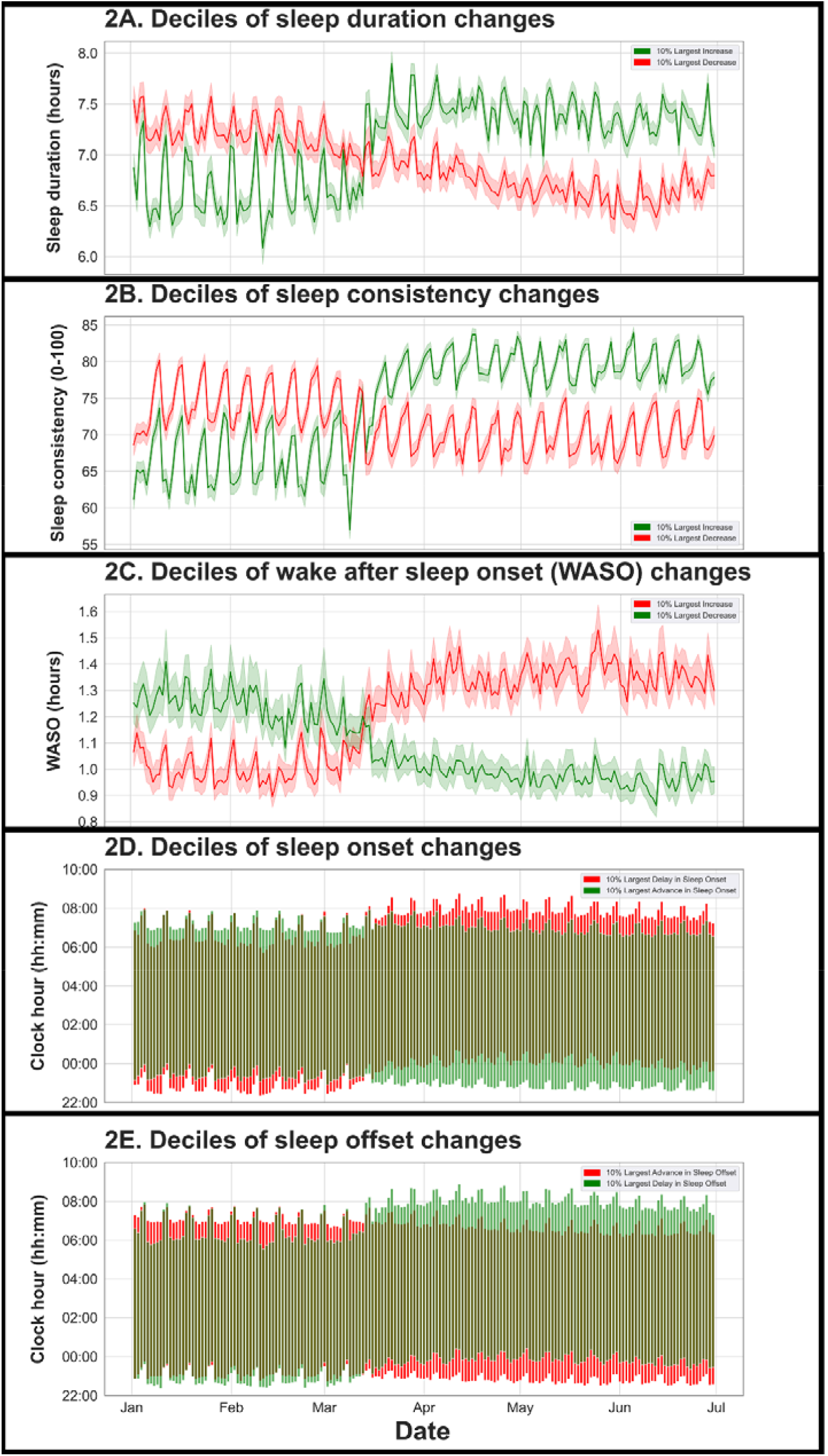
Heterogeneity in changes to sleep duration, consistency, and timing.

Of the 3,845 participants who completed the PHQ-4, 755 (19.6%) screened positive for anxiety or depression symptoms, 1,299 (33.8%) screened positive for COVID-19 trauma- and stressor-related disorder (TSRD) symptoms, 1,208 (32.4%) screened positive for burnout symptoms, and 856 (22.4%) reported that they had started or increased substance use to cope with stress or emotions related to the pandemic (Table S2).

Multivariable analysis including demographic variables and sleep (during both the pre-pandemic, and acute pandemic onset, and early chromic pandemic intervals) or lifestyle variables (i.e., physical activity, alcohol intake) revealed that sleep duration and consistency, along with physical activity, were associated with differences in odds of anxiety or depression symptoms, onset or increased substance use, and burnout symptoms (Table). Regarding sleep duration, compared with participants who slept >7 h in both intervals, individuals who slept <6 h in both intervals had higher odds of anxiety or depression symptoms (aOR = 1.75, 95% CI = 1.14 to 2.69, p = 0.007) and burnout symptoms (aOR = 1.57, 95% CI = 1.07 to 2.29, p = 0.016), as did those who slept 6 to 7 h and those who experienced a decrease in peri-pandemic sleep duration to <6 h from 6-7 h (e.g., burnout symptoms, aOR = 2.22, 95% CI = 1.32 to 3.71, p = 0.001).

Regarding consistency of sleep timing, compared with participants with sleep consistency >80 in both intervals, individuals with sleep consistency <70 in both intervals had higher odds of all three assessed adverse mental and behavioral health symptoms (e.g., onset or increased substance use, aOR = 2.17, 95% CI = 1.48 to 3.19, p < 0.0001). Odds of new or increased substance use were also higher among individuals with sleep consistency of 70-80 during both intervals (aOR = 1.46, 95% CI = 1.06 to 2.01, p = 0.016), and odds of anxiety or depression symptoms were higher among individuals whose sleep consistency decreased from 70-80 in the pre-pandemic interval to <70 in the early chronic pandemic interval (aOR = 2.07, 95% CI = 1.17 to 3.67, p = 0.0009). Odds of adverse mental or behavioral health symptoms were not higher for individuals with decreases in sleep duration or sleep consistency who had optimal duration (>7 h) or consistency (>80) in the pre-pandemic interval, regardless of the magnitude of peri-pandemic decrease.

## Discussion

Among nearly 5,000 active users of a validated sleep wearable with data preceding the onset of the COVID-19 pandemic, we found an acute increase in mean sleep duration and delay in sleep timing in the month when stringent mitigation policies were initially implemented widely across the United States, consistent with literature from the U.S. and around the globe (1-12). Using a novel metric to quantify the consistency of sleep timing modified from the Sleep Regularity Index (30) to account for recency in the weights of comparator sleep episodes, we also found an abrupt and sustained increase in sleep consistency during the pandemic. Across the sample, the magnitude of the increase in mean sleep duration decreased gradually in the subsequent two months, as the mean sleep offset returned to near pre-pandemic times, while the delay in sleep onset persisted. Adverse mental and behavioral health symptoms, including anxiety and depression symptoms; onset, or increased substance use to cope with stress or emotions; and burnout symptoms were associated with pre-pandemic sleep deficiency and inconsistent sleep, but not acute decreases in sleep duration or sleep consistency experienced during the pandemic. These findings indicate that recent past sleep-wake behavior was associated with mental health resilience in response to profound lifestyle changes, such as the stringent social and behavioral interventions (e.g., stay-at-home orders, work-from-home directives) implemented to reduce SARS-CoV-2 transmission during the COVID-19 pandemic.

Analysis of participants with high-magnitude changes to sleep measures revealed disparate changes to sleep during the pandemic, with some participants decreasing in sleep duration and consistency at the onset of the pandemic when there were increases in these measures, on average. Interestingly, whereas participants with substantial increases in these sleep measures exhibited sudden changes immediately following the global pandemic declaration, individuals with substantial decreases exhibited more gradual and persistent decreases. The more consistent trajectory and lack of response to the pandemic declaration suggest that these changes may have contributing factors independent of the pandemic and its mitigation. Given the association between chronic insufficient (25, 35) or inconsistent sleep (25-28) and adverse mental and behavioral health symptoms, differences in changes to sleep during the pandemic may have implications for mental health and are particularly relevant for identifying potential targets for modifiable risk factors. Adjusting for demographic and other lifestyle variables, participants with shorter pre-pandemic sleep duration or lower pre-pandemic consistency of sleep timing had higher odds of adverse mental health symptoms during the early chronic phase of the COVID-19 pandemic compared to participants who attained the recommended sleep duration (>7 h (36)) or high sleep consistency during both intervals. In contrast, participants who had obtained the recommended sleep duration pre-pandemic and participants who had recorded >80 sleep consistency pre-pandemic did not have higher odds of adverse mental health symptoms compared if these sleep measures declined during the pandemic. Together, these results suggest that pre-pandemic sleep duration and consistency may have provided participants with mental health resilience during the initial phase of the pandemic, even among those who experienced worsened sleep during this time.

These findings suggest that sleep could be a modifiable risk factor (1) to be targeted by behavioral interventions designed to enhance mental health resilience (37). At a time when the prevalence of adverse mental and behavioral health symptoms among U.S. adults has increased several-fold (13-15, 19, 38, 39), and non-modifiable risk factors with potential long-term impacts have been introduced (40), the value of modifiable mental health risk factors is of critical importance. Insufficient sleep duration and inconsistent sleep timing are highly prevalent in modern society (41). Alongside many undesirable changes during the COVID-19 pandemic has been a unique opportunity for some to improve sleep behaviors, as demonstrated in our and other studies (1-6, 8-12). Our unique dataset linking mental health and objective high-resolution pre-pandemic sleep-wake data enhances our understanding of the relationships between sleep behaviors and mental health (42), including before, during and after the onset of the COVID-19 pandemic (18). Importantly, there is evidence supporting the efficacy of cognitive and behavioral interventions to improve sleep in adults without sleep disorders (43), providing a precedent for effective measures, including for improvement of sleep to enhance mental health (44). Furthermore, improving sleep may have additional benefits for other major aspects of health, including immune function, cardiovascular health, and diabetes (42), as well as cognitive function (45).

Strengths of this study include the use of objective sleep measures, inclusion of pre-pandemic comparator sleep data, large sample size, psychometrically validated mental health screening instruments, and inclusion of demographic and lifestyle-related variables (i.e., physical activity, alcohol consumption) in multivariable models assessing associations with a comprehensive set of sleep variables (i.e., duration, timing, consistency) (46, 47). Limitations of this study include a lack of pre-pandemic comparator mental health data, non-random recruitment methods and potential seasonal influences on sleep and mood. Regarding the first, the cross-sectional mental health measures preclude a causal interpretation of findings related to mental health. Regarding the second, most sample participants were male, highly educated, employed, and reported higher than the national average household income. Given that income was highly predictive of changes in mobility during the pandemic, with wealthy areas exhibiting larger mobility reductions (48), effects on sleep of stay-at-home orders may be overrepresented in this sample. Moreover, given that WHOOP is a subscription tracker of sleep and fitness, participants may have been more knowledgeable about and motivated to pursue optimal sleep health and fitness than the general United States adult population, which could limit the generalizability of these findings. However, most of the demographic differences in adverse mental health symptom prevalence (e.g., by gender, age, and diurnal preference) were consistent with evidence from the general population (13-15, 19). Finally, it is possible that sleep and mental health responses to the onset of a pandemic may vary with season and be influenced by daylight savings time changes; however, 2019 and 2020 data on time in bed and sleep timing from Capodilupo and Miller (2021) indicate that the magnitude of changes to sleep-wake behavior observed in the months after the COVID-19 pandemic were not observed the year before (12). For example, in 2019, time in bed was slightly shorter during March 10 through May 15 compared to January 1 through March 9, 2019 (by 0.05 ± 0.003 h), and sleep offset time did not differ significantly between the intervals. Comparing the same intervals in 2020, time in bed was considerably longer during March 10 through May 15 (by 0.24 ± 0.003 h), and sleep offset was significantly later (by 29 ± 1 m) (12).

As policymakers grapple with decisions about balancing stringent mitigation measures during future waves of SARS-CoV-2 or other pathogens and mental health, communities, healthcare providers, and public health officials should consider the potential roles of sleep and circadian rhythms in mitigating potential mental health consequences. Future research should evaluate public health programs that include sleep and circadian health as a primary prevention strategy for adverse mental health outcomes and to improve resilience to acute stressors. Sleep duration, timing, and consistency may be modifiable risk factors for adverse mental health in response to stressful life events.

## Materials and Methods

### Study Design and Participant Details

WHOOP users aged ≥18 years with residence in the United States who had recorded the prior seven nocturnal sleep episodes were invited to participate in Internet-based surveys during June 24 to 30, 2020. Participants provided informed electronic consent prior to commencement of the survey. Survey questions were used to determine age- and residence-based eligibility. WHOOP users agreed to allow their deidentified WHOOP wearable data to be used for research purposes, as outlined in the WHOOP Terms and Conditions document. Investigators received anonymized responses, and survey responses were linked with WHOOP wearable data using unique Study Identification codes. The Monash University Human Research Ethics Committee (Melbourne, Victoria, Australia) approved the study protocol.

### WHOOP measures

For this analysis, objective WHOOP variables included sleep duration in hours over 24-hour intervals (calculated as the sum of nocturnal sleep episodes plus nap sleep episodes), sleep consistency (a proprietary metric of the WHOOP platform adapted from the Sleep Regularity Index (30) for daily use by more accounting for recency in weighting comparison sleep-wake episodes), wake after sleep onset (WASO) (calculated as the difference between time in bed and time asleep), and sleep onset and offset. Studies have demonstrated the validity of WHOOP data against polysomnography for detecting sleep-wake determinations (33). Compared to polysomnography, total sleep time estimated by WHOOP did not differ significantly (mean WHOOP total sleep time = 358.7 m ± 98.5 m, mean polysomnography total sleep time = 350.4 m ± 105.2 m, mean difference = 8.2 m ± 32.9 m, p = 0.54). WHOOP sleep-wake data also demonstrated high levels of agreement with polysomnography (89%) and sensitivity to sleep (89% and 95%, respectively) and moderate specificity for wake and Cohen’s kappa for agreement corrected for agreements with polysomnography due to chance (51% and 0.49, respectively) (33).

In addition to sleep measures, upon registration, WHOOP users also input their sex (male or female) and age in years. Ages were categorized as 18-29, 30-44, 45-65, or 65-plus years.

### Survey instrument measures

The survey instrument was developed as part of The COVID-19 Outbreak Public Evaluation (COPE) Initiative (www.thecopeinitiative.org). The survey had been administered to adults in the United States and Australia to assess public attitudes, behaviors, and beliefs about the COVID-19 pandemic and its mitigation, and to assess mental and behavioral health during the pandemic. Demographic variables collected in the survey and included in this analysis were race and ethnicity, U.S. Census region, 2019 household income in United States Dollars, highest education attainment, employment status, unpaid caregiver of adults, political ideology, and diurnal preference assessed using Item 19 of the Horne-Östberg morningness-eveningness questionnaire.

Mental health measures in the survey and included in this analysis were anxiety and depression symptoms assessed using the 4-item Patient Health Questionnaire (PHQ-4), burnout symptoms assessed using the single-item Mini-Z burnout measures, and past-month new or increased substance use to cope with stress or emotions. For the PHQ-4, participants who scored ≥3 out of 6 on the Generalized Anxiety Disorder (GAD-2) and Patient Health Questionnaire (PHQ-2) subscales were considered symptomatic for anxiety or depression, respectively (34). For the Mini-Z, participants who scored ≥3 out of 5 were considered symptomatic for the emotional exhaustion dimension of burnout symptoms (49). Substance use was defined as use of “alcohol, legal or illegal drugs, or prescriptions drugs that are taken in a way not recommended by your doctor.” Participants were asked, “Have you started or increased using substances to help you cope with stress or emotions during the COVID-19 pandemicã” Additional measures included weekly days of physical activity and alcoholic beverage consumption. Physical activity was assessed using a validated single-item physical activity measure (50), “In the past week, on how many days have you done a total of 30 minutes or more of physical activity, which was enough to raise your breathing rateã Physical activity may include sport, exercise, and brisk walking or cycling for recreation or to get to and from places but should not include housework or physical activity that may be part of your job.” Weekly alcoholic beverage consumption was analyzed by multiplying 7 by the answer to the following question: “How many alcoholic beverages did you consume on a typical day in the past weekã”

### Quantification and Statistical Analysis

Study intervals were set as pre-pandemic (1 January to 12 March 2020), acute pandemic onset (13 March to 12 April 2020), and early chronic pandemic (13 April to 30 June 2020). Participants with WHOOP data for ≥70% of nocturnal sleep episodes during each of these intervals were included in the primary analytic sample. Participants who completed the primary adverse mental health symptom screening instrument, the PHQ-4, were included in the mental health analytic subsample for which associations between sleep measures and adverse mental health symptoms were assessed. Chi-square tests were used to assess for demographic differences between participants who did versus did not complete the PHQ-4, with Bonferroni adjustments (n = 9) applied to p values to account for multiple comparisons.

Means and standard deviations were calculated for each WHOOP variable for participants, overall and during each of the survey intervals. Paired t-tests were used to test for differences in mean values for sleep measures between the (1) pre-pandemic and acute pandemic onset and (2) pre-pandemic and early chronic pandemic intervals. Bonferroni adjustments (n = 10) applied to p values and 95% confidence intervals were estimated at the 99.5% confidence level to account for multiple comparisons.

Means and standard deviations were also calculated for each WHOOP variable for deciles of participants (n = 491) with the highest-magnitude changes in sleep measures comparing the pre-pandemic and pandemic intervals (i.e., combined acute pandemic onset and early chronic pandemic intervals). Among each decile paired t-tests were used to test for differences in mean values between these intervals, with Bonferroni adjustments (n = 10) applied to p values and 95% confidence intervals estimated at the 99.5% confidence level to account for multiple comparisons. Finally, multivariable logistic regression models were used to estimate adjusted odds ratios (aORs) and 95% confidence intervals (CIs) for each of the assessed adverse mental health symptoms (anxiety or depression symptoms, new or onset substance use, burnout symptoms) based on pre-pandemic and early chronic pandemic WHOOP measures for sleep duration and sleep consistency. For this analysis, mean sleep duration during these intervals were categorized as <6 h, 6 to 7 h, or >7 h, and mean sleep consistency was categorized as <70, 70-80, or >80 out of 100. As there were two intervals for each variable, there are 3^2^ = 9 categories per variable (e.g., sleep duration <6 h during both intervals, sleep duration <6 hours during the pre-pandemic interval, 6-7 h during the early chronic pandemic interval, etc.). For these models, the reference groups were having recorded the longest mean duration (i.e., >7 h) and highest sleep consistency (i.e., >80) during both intervals. Sex, age group, race/ethnicity, highest education attainment, U.S. Census region, employment status, unpaid caregiver status, diurnal preference, past-week alcoholic beverage consumption, and past-week days with physical activity were included as covariates. Bonferroni adjustments (n = 2) were applied to p values and 95% confidence intervals estimated at the 97.5% confidence level to account for multiple comparisons. Statistical significance for aOR estimates was set as two-sided p < 0.05.

All calculations were performed in Python version 3.7.8 (Python Software Foundation) and R version 4.0.2 (The R Project for Statistical Computing) using the R survey package version 3.29.

**Table 1.**
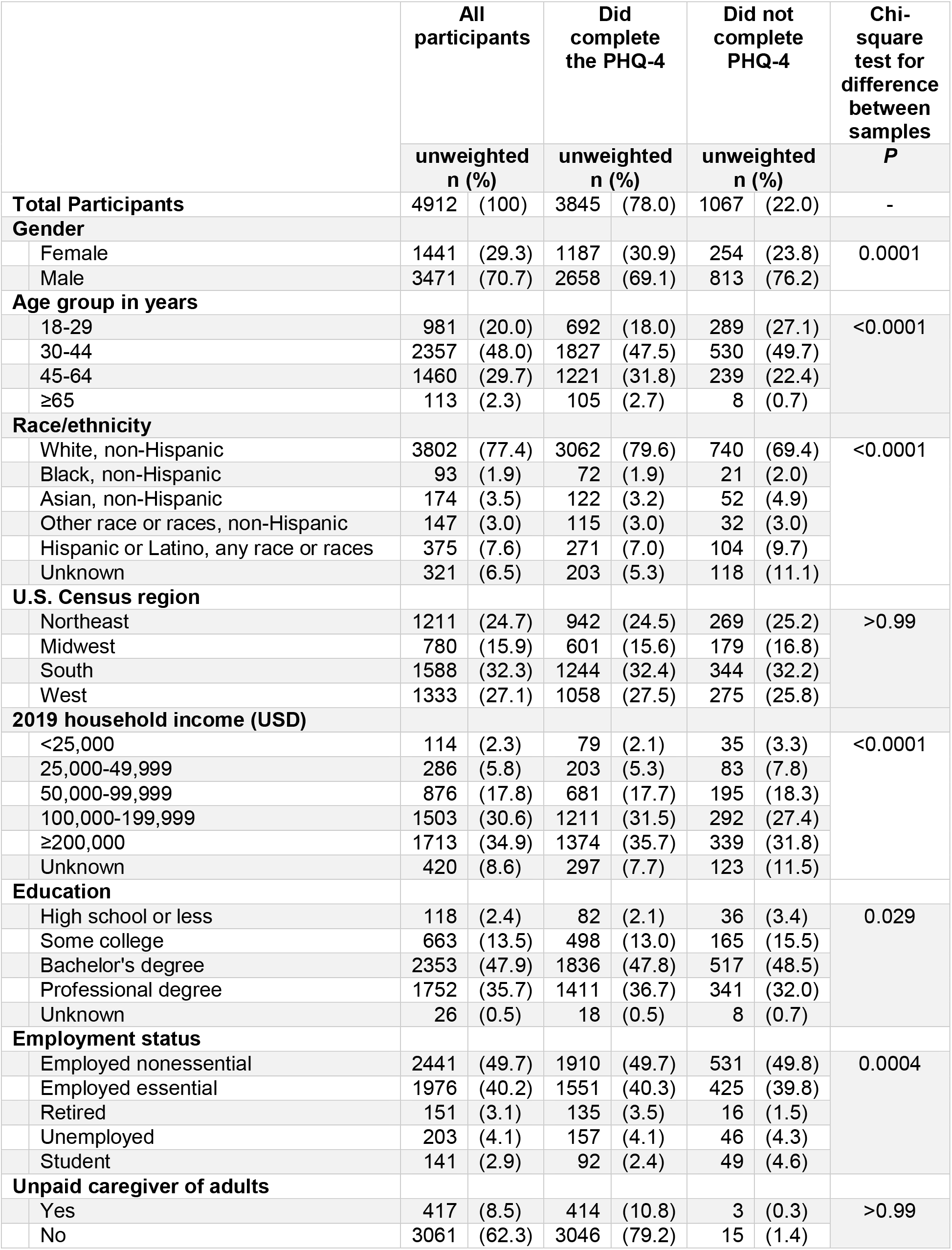
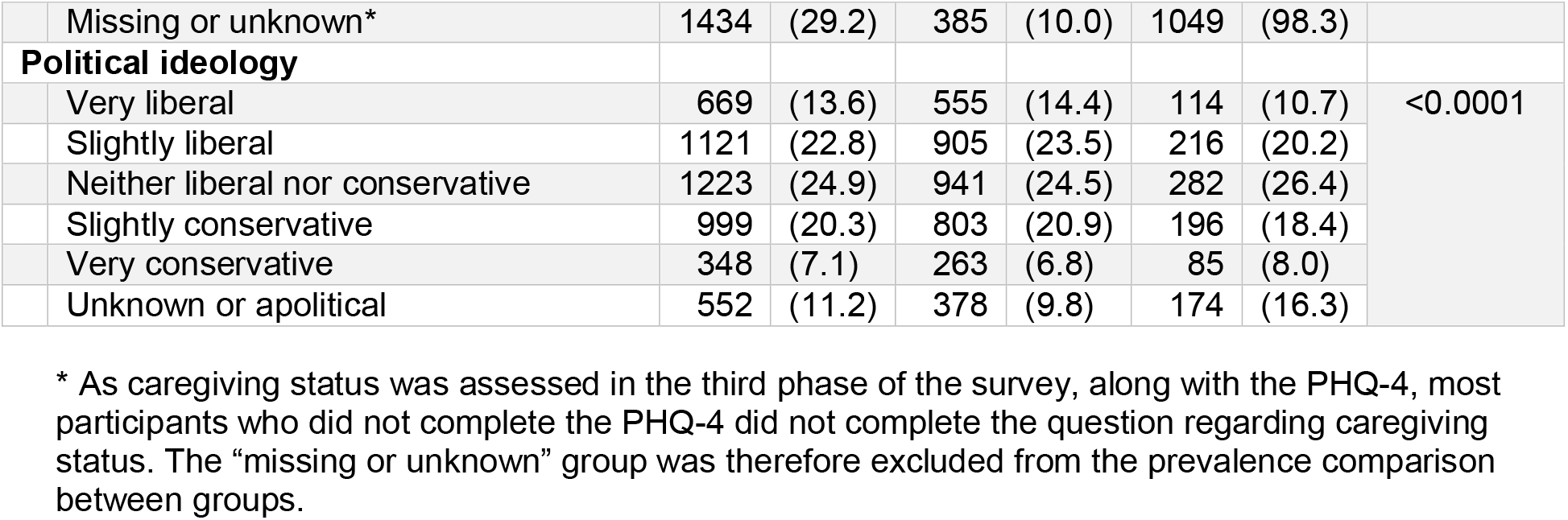
Participant characteristics.

|  | All participants |  | Did complete the PHQ-4 |  | Did not complete PHQ-4 |  | Chi-square test for difference between samples |
| --- | --- | --- | --- | --- | --- | --- | --- |
|  | unweighted n (%) |  | unweighted n (%) |  | unweighted n (%) |  | P |
| **Total Participants** | 4912 | (100) | 3845 | (78.0) | 1067 | (22.0) | - |
| **Gender** |  |  |  |  |  |  |  |
| Female | 1441 | (29.3) | 1187 | (30.9) | 254 | (23.8) | 0.0001 |
| Male | 3471 | (70.7) | 2658 | (69.1) | 813 | (76.2) |  |
| **Age group in years** |  |  |  |  |  |  |  |
| 18-29 | 981 | (20.0) | 692 | (18.0) | 289 | (27.1) | <0.0001 |
| 30-44 | 2357 | (48.0) | 1827 | (47.5) | 530 | (49.7) |  |
| 45-64 | 1460 | (29.7) | 1221 | (31.8) | 239 | (22.4) |  |
| ≥65 | 113 | (2.3) | 105 | (2.7) | 8 | (0.7) |  |
| **Race/ethnicity** |  |  |  |  |  |  |  |
| White, non-Hispanic | 3802 | (77.4) | 3062 | (79.6) | 740 | (69.4) | <0.0001 |
| Black, non-Hispanic | 93 | (1.9) | 72 | (1.9) | 21 | (2.0) |  |
| Asian, non-Hispanic | 174 | (3.5) | 122 | (3.2) | 52 | (4.9) |  |
| Other race or races, non-Hispanic | 147 | (3.0) | 115 | (3.0) | 32 | (3.0) |  |
| Hispanic or Latino, any race or races | 375 | (7.6) | 271 | (7.0) | 104 | (9.7) |  |
| Unknown | 321 | (6.5) | 203 | (5.3) | 118 | (11.1) |  |
| **U.S. Census region** |  |  |  |  |  |  |  |
| Northeast | 1211 | (24.7) | 942 | (24.5) | 269 | (25.2) | >0.99 |
| Midwest | 780 | (15.9) | 601 | (15.6) | 179 | (16.8) |  |
| South | 1588 | (32.3) | 1244 | (32.4) | 344 | (32.2) |  |
| West | 1333 | (27.1) | 1058 | (27.5) | 275 | (25.8) |  |
| **2019 household income (USD)** |  |  |  |  |  |  |  |
| <25,000 | 114 | (2.3) | 79 | (2.1) | 35 | (3.3) | <0.0001 |
| 25,000-49,999 | 286 | (5.8) | 203 | (5.3) | 83 | (7.8) |  |
| 50,000-99,999 | 876 | (17.8) | 681 | (17.7) | 195 | (18.3) |  |
| 100,000-199,999 | 1503 | (30.6) | 1211 | (31.5) | 292 | (27.4) |  |
| ≥200,000 | 1713 | (34.9) | 1374 | (35.7) | 339 | (31.8) |  |
| Unknown | 420 | (8.6) | 297 | (7.7) | 123 | (11.5) |  |
| **Education** |  |  |  |  |  |  |  |
| High school or less | 118 | (2.4) | 82 | (2.1) | 36 | (3.4) | 0.029 |
| Some college | 663 | (13.5) | 498 | (13.0) | 165 | (15.5) |  |
| Bachelor's degree | 2353 | (47.9) | 1836 | (47.8) | 517 | (48.5) |  |
| Professional degree | 1752 | (35.7) | 1411 | (36.7) | 341 | (32.0) |  |
| Unknown | 26 | (0.5) | 18 | (0.5) | 8 | (0.7) |  |
| **Employment status** |  |  |  |  |  |  |  |
| Employed nonessential | 2441 | (49.7) | 1910 | (49.7) | 531 | (49.8) | 0.0004 |
| Employed essential | 1976 | (40.2) | 1551 | (40.3) | 425 | (39.8) |  |
| Retired | 151 | (3.1) | 135 | (3.5) | 16 | (1.5) |  |
| Unemployed | 203 | (4.1) | 157 | (4.1) | 46 | (4.3) |  |
| Student | 141 | (2.9) | 92 | (2.4) | 49 | (4.6) |  |
| **Unpaid caregiver of adults** |  |  |  |  |  |  |  |
| Yes | 417 | (8.5) | 414 | (10.8) | 3 | (0.3) | >0.99 |
| No | 3061 | (62.3) | 3046 | (79.2) | 15 | (1.4) |  |

|  |  |  |  |  |  |  |  |  |
| --- | --- | --- | --- | --- | --- | --- | --- | --- |
|  | Missing or unknown* | 1434 | (29.2) | 385 | (10.0) | 1049 | (98.3) |  |
|  | **Political ideology** |  |  |  |  |  |  |  |
|  | Very liberal | 669 | (13.6) | 555 | (14.4) | 114 | (10.7) | <0.0001 |
|  | Slightly liberal | 1121 | (22.8) | 905 | (23.5) | 216 | (20.2) |  |
|  | Neither liberal nor conservative | 1223 | (24.9) | 941 | (24.5) | 282 | (26.4) |  |
|  | Slightly conservative | 999 | (20.3) | 803 | (20.9) | 196 | (18.4) |  |
|  | Very conservative | 348 | (7.1) | 263 | (6.8) | 85 | (8.0) |  |
|  | Unknown or apolitical | 552 | (11.2) | 378 | (9.8) | 174 | (16.3) |  |

\* As caregiving status was assessed in the third phase of the survey, along with the PHQ-4, most participants who did not complete the PHQ-4 did not complete the question regarding caregiving status. The “missing or unknown” group was therefore excluded from the prevalence comparison between groups.

**Table 2.** Adjusted odds ratios (aORs) for adverse mental health symptoms by pre- and per-pandemic sleep characteristics.

|  | Anxiety or depression symptoms (N=3845) |  | Started or increased substance use (N=3820) |  | Burnout symptoms (N=3734) |  |
| --- | --- | --- | --- | --- | --- | --- |
|  | aOR (95% CI) | P | aOR (95% CI) | P | aOR (95% CI) | P |
| **Sleep duration (reference: >7 hours before and during the pandemic)** |  |  |  |  |  |  |
| Both <6 h | 1.75 (1.14, 2.69) | 0.007 | 1.11 (0.69, 1.78) | >0.99 | 1.57 (1.07, 2.29) | 0.016 |
| <6 h to 6-7 h | 1.12 (0.64, 1.98) | >0.99 | 1.21 (0.71, 2.04) | 0.85 | 1.22 (0.77, 1.93) | 0.66 |
| <6 h to >7 h | NO ESTIMATE | - | 2.20 (0.33, 14.55) | 0.70 | 0.82 (0.07, 9.88) | >0.99 |
| Both 6-7 h | 1.30 (1.03, 1.65) | 0.025 | 1.21 (0.96, 1.52) | 0.13 | 1.39 (1.13, 1.70) | 0.001 |
| 6-7 h to <6 h | 1.96 (1.09, 3.54) | 0.021 | 1.19 (0.65, 2.17) | >0.99 | 2.22 (1.32, 3.71) | 0.001 |
| 6-7 h to >7 h | 1.23 (0.90, 1.67) | 0.27 | 0.96 (0.70, 1.31) | >0.99 | 1.22 (0.93, 1.60) | 0.19 |
| >7 h to <6 h | 0.86 (0.06, 13.38) | >0.99 | 4.88 (0.34, 69.45) | 0.36 | NO ESTIMATE | - |
| >7 h to 6-7 h | 0.94 (0.60, 1.46) | >0.99 | 1.08 (0.72, 1.64) | >0.99 | 0.86 (0.59, 1.26) | 0.75 |
| **Sleep consistency (reference: >80 out of 100 before and during the pandemic)** |  |  |  |  |  |  |
| Both <70 | 1.74 (1.19, 2.55) | 0.002 | 2.17 (1.48, 3.19) | <0.001 | 1.77 (1.28, 2.45) | <0.001 |
| <70 to 70-80 | 1.38 (0.95, 1.99) | 0.10 | 1.38 (0.95, 1.99) | 0.10 | 1.27 (0.94, 1.73) | 0.16 |
| <70 to >80 | 1.13 (0.57, 2.25) | >0.99 | 1.35 (0.69, 2.65) | 0.64 | 0.88 (0.48, 1.62) | >0.99 |
| Both 70-80 | 1.34 (0.97, 1.85) | 0.088 | 1.46 (1.06, 2.01) | 0.016 | 1.22 (0.93, 1.60) | 0.19 |
| 70-80 to <70 | 2.07 (1.17, 3.67) | 0.009 | 1.66 (0.91, 3.03) | 0.12 | 1.08 (0.63, 1.84) | >0.99 |
| 70-80 to >80 | 1.11 (0.79, 1.56) | 0.95 | 1.17 (0.84, 1.63) | 0.60 | 0.99 (0.75, 1.31) | >0.99 |
| >80 to <70 | 3.41 (0.16, 74.07) | 0.74 | 2.14 (0.07, 66.56) | >0.99 | NO ESTIMATE | - |
| >80 to 70-80 | 1.35 (0.66, 2.75) | 0.70 | 1.62 (0.80, 3.30) | 0.25 | 1.31 (0.71, 2.42) | 0.65 |

## Data Availability

Public data sharing for these purposes is ethically and legally restricted. The data is collected and used with the consent of individuals who purchase a WHOOP membership (collectively, "WHOOP Members") and agree to the WHOOP terms of use (https://www.whoop.com/termsofuse/) and privacy policy (https://www.whoop.com/privacy/full-privacy-policy/). While WHOOP terms of use and privacy policy permit WHOOP to use collected data that has been aggregated or de-identified in a manner that no longer identifies an individual for informational purposes, analytics or WHOOP's own research purposes, the consent obtained from WHOOP Members does not extend to making the data publicly available for a third party to use for its own purposes. As such, WHOOP's legal department will not permit the data to be shared for these purposes. Data access queries can be directed to the WHOOP Executive team via the Vice President of Performance at WHOOP - Kristen Holmes.

## Acknowledgments

The authors thank Thomas Rand (WHOOP, Inc.) for coding the survey onto the WHOOP interface, as well as Rebecca Robbins, Ph.D. (Brigham and Women’s Hospital, Harvard Medical School), Laura K. Barger, Ph.D. (Brigham and Women’s Hospital, Harvard Medical School), and Elise R. Facer-Childs, Ph.D. (Monash University) for their contributions to the initial survey instrument for The COPE Initiative. M.É.C. gratefully acknowledges funding by The Kinghorn Foundation through a 2020 to 2021 Australian-American Fulbright Scholarship.

## Author Contributions

Mark É. Czeisler: Conceptualization, Data curation, Formal Analysis, Funding acquisition, Methodology, Visualization, Writing – original draft, Writing – review & editing: Emily R. Capodilupo: Conceptualization, Data curation, Formal Analysis, Funding acquisition, Methodology, Visualization, Writing – review & editing: Matthew D. Weaver: Conceptualization, Formal Analysis, Visualization, Writing – review & editing: Charles A. Czeisler: Conceptualization, Funding acquisition, Methodology, Writing – review & editing: Mark E. Howard: Conceptualization, Funding acquisition, Methodology, Supervision, Writing – review & editing: Shantha M.W. Rajaratnam: Conceptualization, Formal Analysis, Funding acquisition, Methodology, Supervision, Visualization, Writing – review & editing

## Supplementary Information for

**Fig. S1.**
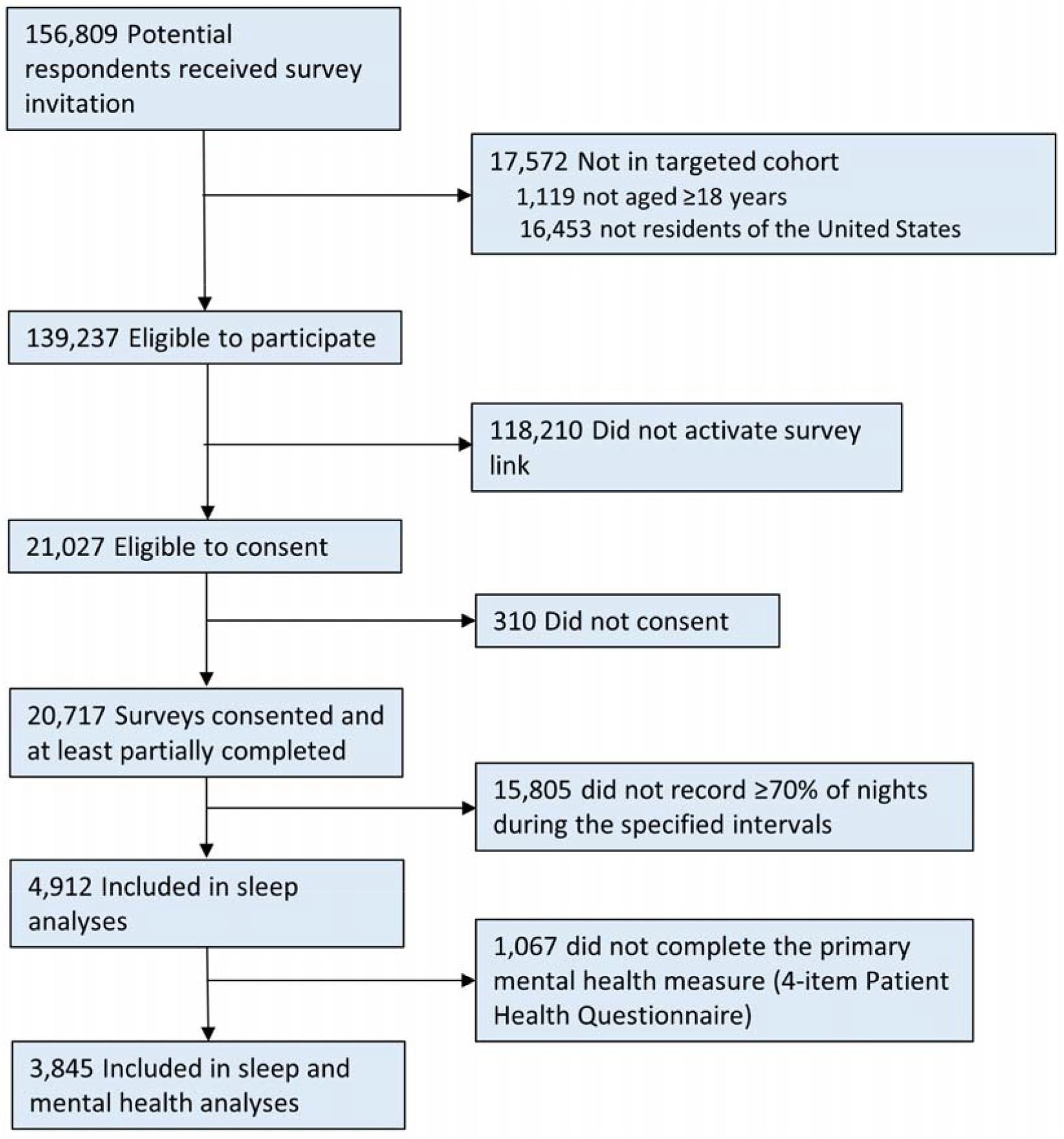
Flow of the survey respondents.

**Table S1.** Adverse mental health symptoms and sleep characteristics of the respondents.

|  |  | All respondents |  |  | Did complete the PHQ-4 |  |  |
| --- | --- | --- | --- | --- | --- | --- | --- |
| MENTAL HEALTH |  |  |  |  |  |  |  |
|  | Symptoms of anxiety or depressive disorder (N=3845)–n (%) | - | - | - | 755 | (19.6) | - |
|  | Started or increased substance use (N=3820)–n (%) | - | - | - | 856 | (22.4) | - |
|  | Symptoms of burnout (N=3734)–n (%) | - | - | - | 1208 | (32.4) | - |
| SLEEP |  |  |  | p |  |  | p |
| Sleep duration in hrs–mean (sd) |  |  |  |  |  |  |  |
|  | Pre-pandemic | 6.95 | (0.687) | - | 6.96 | (0.687) | - |
|  | Acute pandemic | 7.20 | (0.751) | - | 7.21 | (0.75) | - |
|  | Early chronic pandemic | 7.04 | (0.705) | - | 7.06 | (0.702) | - |
|  | Difference: Acute vs Pre (95% CI) | 0.25 | (0.237, 0.27) | <0.0001 | 0.25 | (0.231, 0.269) | <0.0001 |
|  | Difference: Early chronic vs Pre (95% CI) | 0.09 | (0.076, 0.107) | <0.0001 | 0.09 | (0.077, 0.112) | <0.0001 |
| Sleep consistency–mean (sd) |  |  |  |  |  |  |  |
|  | Pre-pandemic | 73.30 | (7.179) | - | 73.53 | (7.222) | - |
|  | Acute pandemic | 76.81 | (7.705) | - | 77.00 | (7.78) | - |
|  | Early chronic pandemic | 77.36 | (7.259) | - | 77.59 | (7.33) | - |
|  | Difference: Acute vs Pre (95% CI) | 3.51 | (3.295, 3.728) | <0.0001 | 3.47 | (3.226, 3.715) | <0.0001 |
|  | Difference: Early chronic vs Pre (95% CI) | 4.06 | (3.856, 4.267) | <0.0001 | 4.07 | (3.834, 4.297) | <0.0001 |
| Wake after sleep onset in hrs–mean (sd) |  |  |  |  |  |  |  |
|  | Pre-pandemic | 1.15 | (0.576) | - | 1.14 | (0.527) | - |
|  | Acute pandemic | 1.10 | (0.487) | - | 1.10 | (0.494) | - |
|  | Early chronic pandemic | 1.16 | (0.662) | - | 1.15 | (0.668) | - |
|  | Difference: Acute vs Pre (95% CI) | -0.05 | (-0.074, -0.031) | <0.0001 | -0.05 | (-0.067, -0.025) | <0.0001 |
|  | Difference: Early chronic vs Pre (95% CI) | 0.01 | (-0.022, 0.035) | >0.99 | 0.01 | (-0.02, 0.039) | >0.99 |
| Sleep onset–mean (sd) |  |  |  |  |  |  |  |
|  | Pre-pandemic | 22:50 | (70.360) | - | 22:48 | (70.400) | - |
|  | Acute pandemic | 23:09 | (77.306) | - | 23:06 | (76.401) | - |
|  | Early chronic pandemic | 23:08 | (75.475) | - | 23:05 | (74.669) | - |
|  | Difference: Acute vs Pre (95% CI) | 0:18 | (17.378, 20.045) | <0.0001 | 0:18 | (16.663, 19.626) | <0.0001 |
|  | Difference: Early chronic vs Pre (95% CI) | 0:17 | (16.470, 19.289) | <0.0001 | 0:17 | (16.029, 19.132) | <0.0001 |

| **Sleep offset–mean (sd)** |  |  |  |  |  |  |
| --- | --- | --- | --- | --- | --- | --- |
| Pre-pandemic | 6:42 | (69.723) | - | 6:41 | (69.670) | - |
| Acute pandemic | 7:19 | (77.076) | - | 7:16 | (76.566) | - |
| Early chronic pandemic | 7:07 | (75.559) | - | 7:05 | (74.840) | - |
| Difference: Acute vs Pre (95% CI) | 0:36 | (35.111, 38.106) | <0.0001 | 0:35 | (33.997, 37.346) | <0.0001 |
| Difference: Early chronic vs Pre (95% CI) | 0:25 | (23.629, 26.714) | <0.0001 | 0:24 | (23.234, 26.669) | <0.0001 |

**Table S2.**
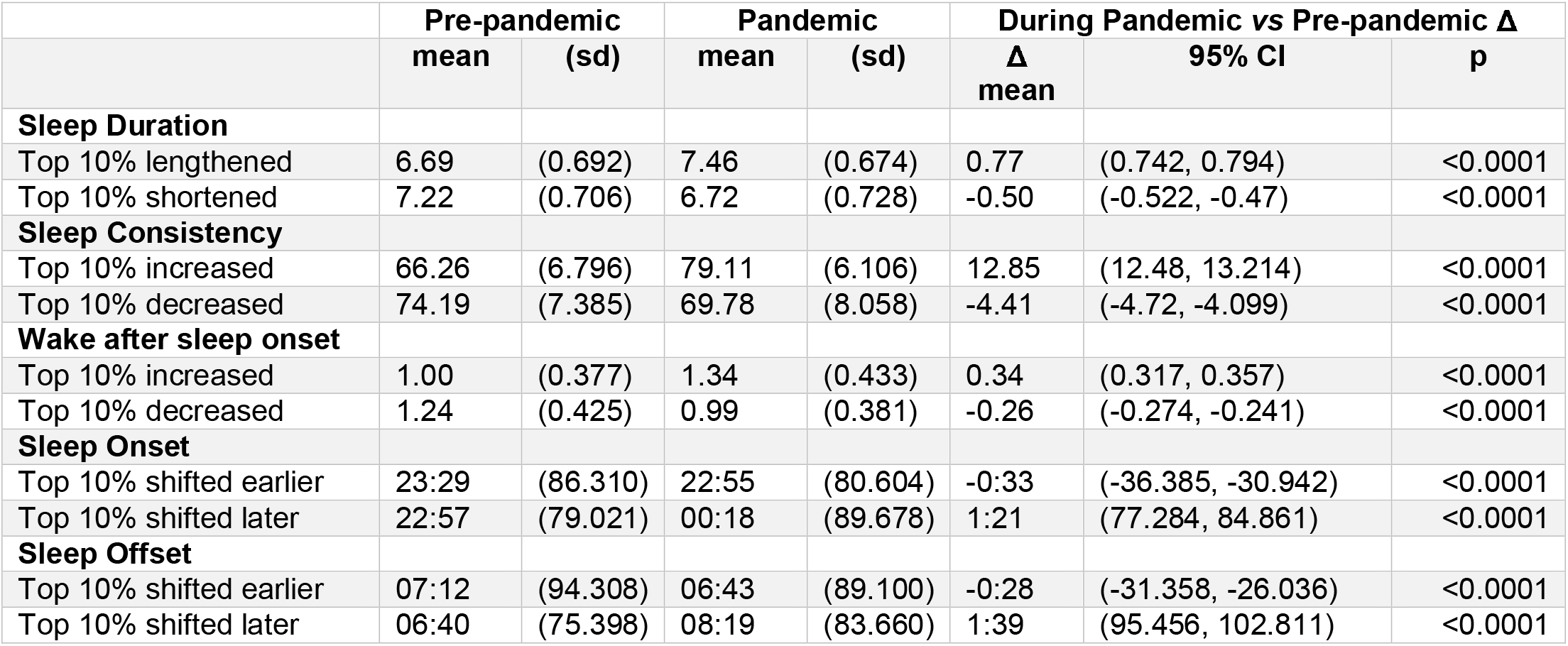
Varying trends in sleep measures during the COVID-19 pandemic.

## References

1. K. P. Wright, Jr. et al., Sleep in university students prior to and during COVID-19 Stay-at-Home orders. Curr Biol 30, R797–R798 (2020).

2. C. Gao, M. K. Scullin, Sleep health early in the coronavirus disease 2019 (COVID-19) outbreak in the United States: integrating longitudinal, cross-sectional, and retrospective recall data. Sleep Med 73, 1–10 (2020).

3. M. J. Leone, M. Sigman, D. A. Golombek, Effects of lockdown on human sleep and chronotype during the COVID-19 pandemic. Curr Biol 30, R930–R931 (2020).

4. M. Sinha, B. Pande, R. Sinha, Impact of COVID-19 lockdown on sleep-wake schedule and associated lifestyle related behavior: A national survey. J Public Health Res 9, 1826 (2020).

5. E. R. Facer-Childs, D. Hoffman, J. N. Tran, S. P. A. Drummond, S. M. W. Rajaratnam, Sleep and mental health in athletes during COVID-19 lockdown. Sleep 10.1093/sleep/zsaa261 (2021).

6. F. D. Genta et al., COVID-19 pandemic impact on sleep habits, chronotype and health-related quality of life among high school students: a longitudinal study. J Clin Sleep Med 10.5664/jcsm.9196 (2021).

7. V. Alfonsi et al., COVID-19 lockdown and poor sleep quality: Not the whole story. J Sleep Res 10.1111/jsr.13368, e13368 (2021).

8. R. Robbins et al., Estimated Sleep Duration Before and During the COVID-19 Pandemic in Major Metropolitan Areas on Different Continents: Observational Study of Smartphone App Data. J Med Internet Res 23, e20546 (2021).

9. J. L. Pepin et al., Greatest changes in objective sleep architecture during COVID-19 lockdown in night-owls with increased REM sleep. Sleep 10.1093/sleep/zsab075 (2021).

10. N. Rezaei, M. A. Grandner, Changes in sleep duration, timing, and variability during the COVID-19 pandemic: Large-scale Fitbit data from 6 major US cities. Sleep Health 10.1016/j.sleh.2021.02.008 (2021).

11. J. L. Ong et al., COVID-19-related mobility reduction: heterogenous effects on sleep and physical activity rhythms. Sleep 44 (2021).

12. E. R. Capodilupo, D. J. Miller, Changes in health promoting behavior during COVID-19 physical distancing: Utilizing WHOOP data to Examine Trends in Sleep, Activity, and Cardiovascular Health. medRxiv https://doi.org/10.1101/2020.06.07.20124685 (2020).

13. C. K. Ettman et al., Prevalence of Depression Symptoms in US Adults Before and During the COVID-19 Pandemic. JAMA Netw Open 3, e2019686 (2020).

14. M. E. Czeisler et al., Early public adherence with and support for stay-at-home COVID-19 mitigation strategies despite adverse life impact: a transnational cross-sectional survey study in the United States and Australia. BMC Public Health 21, 503 (2021).

15. M. E. Czeisler et al., Mental Health, Substance Use, and Suicidal Ideation During the COVID-19 Pandemic - United States, June 24-30, 2020. MMWR Morb Mortal Wkly Rep 69, 1049–1057 (2020).

16. P. K. Alvaro, R. M. Roberts, J. K. Harris, A Systematic Review Assessing Bidirectionality between Sleep Disturbances, Anxiety, and Depression. Sleep 36, 1059–1068 (2013).

17. E. A. Holmes et al., Multidisciplinary research priorities for the COVID-19 pandemic: a call for action for mental health science. Lancet Psychiatry 7, 547–560 (2020).

18. P. Varma, M. Junge, H. Meaklim, M. L. Jackson, Younger people are more vulnerable to stress, anxiety and depression during COVID-19 pandemic: A global cross-sectional survey. Prog Neuropsychopharmacol Biol Psychiatry 109, 110236 (2020).

19. M. E. Czeisler et al., Follow-up Survey of US Adult Reports of Mental Health, Substance Use, and Suicidal Ideation During the COVID-19 Pandemic, September 2020. JAMA Netw Open 4, e2037665 (2021).

20. C. Franceschini et al., Poor Sleep Quality and Its Consequences on Mental Health During the COVID-19 Lockdown in Italy. Front Psychol 11, 574475 (2020).

21. D. S. Lauderdale, K. L. Knutson, L. L. Yan, K. Liu, P. J. Rathouz, Self-reported and measured sleep duration: how similar are theyã Epidemiology 19, 838–845 (2008).

22. C. L. Jackson, S. R. Patel, W. B. Jackson, 2nd, P. L. Lutsey, S. Redline, Agreement between self-reported and objectively measured sleep duration among white, black, Hispanic, and Chinese adults in the United States: Multi-Ethnic Study of Atherosclerosis. Sleep 41 (2018).

23. E. M. Cespedes et al., Comparison of Self-Reported Sleep Duration With Actigraphy: Results From the Hispanic Community Health Study/Study of Latinos Sueno Ancillary Study. Am J Epidemiol 183, 561–573 (2016).

24. D. J. Dijk et al., Sleep, performance, circadian rhythms, and light-dark cycles during two space shuttle flights. Am J Physiol Regul Integr Comp Physiol 281, R1647–1664 (2001).

25. Y. Fang, D. B. Forger, E. Frank, S. Sen, C. Goldstein, Day-to-day variability in sleep parameters and depression risk: a prospective cohort study of training physicians. NPJ Digit Med 4, 28 (2021).

26. L. M. Lyall et al., Association of disrupted circadian rhythmicity with mood disorders, subjective wellbeing, and cognitive function: a cross-sectional study of 91 105 participants from the UK Biobank. Lancet Psychiatry 5, 507–514 (2018).

27. B. Bei, R. Manber, N. B. Allen, J. Trinder, J. F. Wiley, Too Long, Too Short, or Too Variableã Sleep Intraindividual Variability and Its Associations With Perceived Sleep Quality and Mood in Adolescents During Naturalistically Unconstrained Sleep. Sleep 40 (2017).

28. P. H. Finan, P. J. Quartana, M. T. Smith, The Effects of Sleep Continuity Disruption on Positive Mood and Sleep Architecture in Healthy Adults. Sleep 38, 1735–1742 (2015).

29. G. Medic, M. Wille, M. E. Hemels, Short- and long-term health consequences of sleep disruption. Nat Sci Sleep 9, 151–161 (2017).

30. A. J. K. Phillips et al., Irregular sleep/wake patterns are associated with poorer academic performance and delayed circadian and sleep/wake timing. Sci Rep 7, 3216 (2017).

31. J. R. Lunsford-Avery, M. M. Engelhard, A. M. Navar, S. H. Kollins, Validation of the Sleep Regularity Index in Older Adults and Associations with Cardiometabolic Risk. Sci Rep 8, 14158 (2018).

32. S. Berryhill et al., Effect of wearables on sleep in healthy individuals: a randomized crossover trial and validation study. J Clin Sleep Med 16, 775–783 (2020).

33. D. J. Miller et al., A validation study of the WHOOP strap against polysomnography to assess sleep. J Sports Sci 38, 2631–2636 (2020).

34. B. Lowe et al., A 4-item measure of depression and anxiety: validation and standardization of the Patient Health Questionnaire-4 (PHQ-4) in the general population. J Affect Disord 122, 86–95 (2010).

35. V. K. Chattu et al., The Global Problem of Insufficient Sleep and Its Serious Public Health Implications. Healthcare (Basel) 7 (2018).

36. M. Hirshkowitz et al., National Sleep Foundation’s sleep time duration recommendations: methodology and results summary. Sleep Health 1, 40–43 (2015).

37. S. Brand et al., Adolescents with greater mental toughness show higher sleep efficiency, more deep sleep and fewer awakenings after sleep onset. J Adolesc Health 54, 109–113 (2014).

38. A. Vahratian, S. J. Blumberg, E. P. Terlizzi, J. S. Schiller, Symptoms of Anxiety or Depressive Disorder and Use of Mental Health Care Among Adults During the COVID-19 Pandemic - United States, August 2020-February 2021. MMWR Morb Mortal Wkly Rep 70, 490–494 (2021).

39. L. R. McKnight-Eily et al., Racial and Ethnic Disparities in the Prevalence of Stress and Worry, Mental Health Conditions, and Increased Substance Use Among Adults During the COVID-19 Pandemic - United States, April and May 2020. MMWR Morb Mortal Wkly Rep 70, 162–166 (2021).

40. E. J. Raker, M. Zacher, S. R. Lowe, Lessons from Hurricane Katrina for predicting the indirect health consequences of the COVID-19 pandemic. Proc Natl Acad Sci U S A 117, 12595–12597 (2020).

41. Y. Liu et al., Prevalence of Healthy Sleep Duration among Adults--United States, 2014. MMWR Morb Mortal Wkly Rep 65, 137–141 (2016).

42. N. F. Watson et al., Recommended Amount of Sleep for a Healthy Adult: A Joint Consensus Statement of the American Academy of Sleep Medicine and Sleep Research Society. Sleep 38, 843–844 (2015).

43. B. Murawski, L. Wade, R. C. Plotnikoff, D. R. Lubans, M. J. Duncan, A systematic review and meta-analysis of cognitive and behavioral interventions to improve sleep health in adults without sleep disorders. Sleep Med Rev 40, 160–169 (2018).

44. H. Christensen et al., Effectiveness of an online insomnia program (SHUTi) for prevention of depressive episodes (the GoodNight Study): a randomised controlled trial. Lancet Psychiatry 3, 333–341 (2016).

45. S. Diekelmann, Sleep for cognitive enhancement. Front Syst Neurosci 8, 46 (2014).

46. D. J. Buysse, Sleep health: can we define itã Does it matterã Sleep 37, 9–17 (2014).

47. L. Dong, A. J. Martinez, D. J. Buysse, A. G. Harvey, A composite measure of sleep health predicts concurrent mental and physical health outcomes in adolescents prone to eveningness. Sleep Health 5, 166–174 (2019).

48. J. A. Weill, M. Stigler, O. Deschenes, M. R. Springborn, Social distancing responses to COVID-19 emergency declarations strongly differentiated by income. Proc Natl Acad Sci U S A 117, 19658–19660 (2020).

49. E. D. Dolan et al., Using a single item to measure burnout in primary care staff: a psychometric evaluation. J Gen Intern Med 30, 582–587 (2015).

50. K. Milton, F. C. Bull, A. Bauman, Reliability and validity testing of a single-item physical activity measure. Br J Sports Med 45, 203–208 (2011).

